# Epidemiological Analysis of the 2026 Bundibugyo Virus Disease Outbreak and Rapid Risk Assessment for North Africa and Europe

**DOI:** 10.64898/2026.07.19.26358411

**Authors:** Oussama Wail Bouhentala, Mohamed Yazid Kadir

## Abstract

**Background:** In 2026, the Democratic Republic of the Congo (DRC) experienced the largest recorded outbreak of Ebola disease caused by Bundibugyo virus, with epidemiologically linked importations and secondary transmission in Uganda. This study analysed the publicly reported trajectory and assessed the risk of introduction and onward transmission in North Africa and Europe.

**Methods:** Public surveillance reports from the World Health Organization (WHO), European Centre for Disease Prevention and Control (ECDC), Africa CDC, national ministries of health, and peer-reviewed sources were synthesised through 15–17 July 2026. Headline counts and crude case-fatality ratios were restricted to laboratory-confirmed cases. Average notification rates were calculated from cumulative DRC counts. Exact Poisson intervals used the Garwood method, and the June–July rate ratio was estimated on the log scale. Risk was assessed across introduction likelihood, conditional onward-transmission likelihood, impact, and confidence.

**Results:** By 15 July, the DRC had reported 2,124 confirmed cases and 828 deaths (crude confirmed-case fatality ratio, 39.0%) across 46 health zones in five provinces. Uganda had reported 20 confirmed cases and two confirmed deaths: 15 imported infections and five secondary cases, with no documented community transmission. DRC notifications averaged 47.4 per day during 1–15 July versus 35.9 per day during 2–29 June (rate ratio 1.32; counting-model 95% interval 1.20–1.46). WHO reported that more than 80% of new cases were detected outside known contact lists, while 119 confirmed healthcare-worker infections and 36 deaths had occurred. Introduction likelihood was assessed as very low to low for North Africa and very low for the general European population; delayed recognition in routine healthcare was the principal scenario for limited secondary transmission.

**Interpretation:** Available indicators were inconsistent with effective control in eastern DRC at the data cut-off. Public reporting-date series cannot separate transmission from changing ascertainment, but they showed no sustained decline. Preparedness in North Africa and Europe should prioritise complete exposure histories, rapid isolation, validated diagnostics, protected clinical care, and contact management rather than reliance on border screening.

## 1. Introduction

Ebola disease caused by Bundibugyo virus (BDBV) was confirmed in the Democratic Republic of the Congo (DRC) and Uganda in May 2026. The outbreak expanded rapidly in eastern DRC in a setting characterised by insecurity, displacement, high mobility, constrained health services, and repeated occupational exposure. On 17 May 2026, the WHO Director-General determined that the event constituted a Public Health Emergency of International Concern under the International Health Regulations (IHR), while concluding that it did not constitute a pandemic emergency. WHO subsequently issued temporary recommendations for affected countries, neighbouring states, and other States Parties.[3,5,20]

The event is epidemiologically and operationally distinct from outbreaks caused by Ebola virus because no vaccine or therapeutic is licensed specifically for BDBV. Candidate vaccines and therapeutics are under evaluation, and WHO initiated the PARTNERS platform trial in July 2026. The absence of an established BDBV-specific medical countermeasure increases the importance of early detection, high-quality supportive care, infection prevention and control (IPC), contact tracing, community engagement, and safe and dignified burial practices.[21–23,25,34]

International exportation had already occurred by mid-July. Uganda reported imported infections and epidemiologically linked secondary transmission; France detected a returning medical doctor; and two United States humanitarian workers diagnosed in the DRC were medically evacuated to Germany. These events demonstrate that exportation is possible, but observed events remained concentrated among occupationally exposed persons and controlled evacuations rather than ordinary-traveller community spread.[1,3,27]

Assessing risk outside the affected region requires separating two questions that are frequently collapsed: whether an infected person may arrive, and whether an imported case would generate secondary or sustained transmission. This distinction is particularly important for North African and European settings, where introduction is biologically possible but sustained community transmission should remain uncommon if the first case is recognised rapidly and managed with appropriate precautions.

This study aimed to: (1) describe the magnitude, notification trajectory, geographical distribution, and outcome indicators of the outbreak; (2) evaluate limitations of the public surveillance data; (3) identify the principal mechanisms sustaining transmission; (4) assess introduction, onward-transmission, impact, and confidence for North Africa and Europe; and (5) identify operational preparedness priorities, with a specific focus on Algeria.

## 2. Methods

### 2.1 Study design, scope, and evidence sources

This was a rapid public-health analysis of publicly available aggregate surveillance data and official risk assessments. The principal sources were WHO Disease Outbreak News, WHO/IHR statements and technical guidance, ECDC surveillance and modelling reports, WHO regional offices, national ministry-of-health dashboards and updates, and peer-reviewed primary studies for historical BDBV outbreaks, viral persistence, and candidate countermeasures. Airline sources were used only to establish the existence of plausible routes and not to estimate passenger volumes.

The main epidemiological data cut-off was 15 July 2026 for the DRC confirmed-case and death series, and 17 July 2026 for DRC situational indicators and Uganda status. The primary North African scope comprised Algeria, Egypt, Libya, Morocco, Sudan, and Tunisia. Mauritania was included as a Maghreb-adjacent state with regional travel and public-health links. The main qualitative risk horizon was four to eight weeks; narrative planning scenarios extended to three months.

### 2.2 Case definitions and denominator discipline

The main DRC trajectory used laboratory-confirmed cases and deaths among confirmed cases.

Suspected, probable, and confirmed categories were not summed across reports because individuals and deaths can be reclassified as investigations progress. Crude case-fatality ratios (CFRs) were therefore calculated only within explicitly defined confirmed-case denominators and were not interpreted as infection-fatality ratios. Uganda’s confirmed series excluded one probable fatal case, which was described separately.[3,26]

When official sources reported different recovery, hospitalisation, or contact-tracing values, the most clearly defined primary-source figure at the stated cut-off was used. Unresolved discrepancies were retained as data-quality limitations rather than reconciled through undocumented assumptions.

### 2.3 Notification-rate analysis

Cumulative confirmed DRC counts were indexed by reporting date rather than symptom-onset date. Interval-specific average notification rates were calculated as the change in cumulative confirmed cases divided by the number of days between reports. Exact 95% Poisson counting intervals were calculated using the Garwood method. For a count k observed over t days, interval limits for the count were 0.5χ^2^(0.025, 2k) and 0.5χ^2^(0.975, 2[k+1]), divided by t. These intervals describe counting variation only and do not incorporate systematic uncertainty from backlogs, under-ascertainment, changing testing, or reporting delays.

The notification-rate ratio comparing 1–15 July with 2–29 June was calculated as (C_1_/t_1_)/(C_0_/t_0_). The 95% interval was estimated on the log scale using SE[log(RR)] = √(1/C_1_ + 1/C_0_). The interval from 29 June to 1 July was excluded from the aggregate month comparison because it crossed the month boundary.

Arithmetic cumulative doubling times were treated as descriptive properties of the cumulative curve and not as evidence of changes in transmission. No reproduction number was estimated because onset-date line-list data were unavailable.

### 2.4 Risk-assessment framework

Risk was separated into four dimensions: introduction likelihood, conditional onward-transmission likelihood after an importation, impact if detection was delayed, and confidence in the judgement. Categories were comparative reference classes rather than calibrated probabilities. Boundary ratings such as “very low to low” were retained when the evidence did not support a more precise classification. Impact was assessed independently of likelihood, considering expected clinical severity, occupational exposure, contact-tracing burden, service disruption, public concern, and health-system resilience. The ratings were calibrated against WHO’s 6 June assessment, which classified risk as very high in the DRC, high in Uganda and bordering countries, low in the remainder of the WHO African Region, and low globally.[5]

### 2.5 Ethics and reporting

The study used only publicly available aggregate surveillance information and did not involve identifiable participant-level data, recruitment, intervention, or access to confidential records. Formal research-ethics review and participant consent were therefore not applicable. The analysis was designed to be reproducible from the counts and formulas reported in the manuscript and Appendix A.

## 3. Results

### 3.1 Outbreak magnitude and geographical distribution

As of 15 July 2026, the DRC had reported 2,124 laboratory-confirmed cases, 828 deaths among confirmed cases, 390 reported recoveries, and 725 patients hospitalised in isolation. Confirmed cases had been reported from 46 of 140 health zones across five provinces. Thirty-eight affected health zones had reported cases within the previous 21 days. Ituri accounted for 89.6% of confirmed cases and 83.6% of confirmed deaths.[1,3]

The published status totals did not fully reconcile: 828 deaths, 390 recoveries, and 725 hospitalised patients summed to 1,943, leaving 181 of 2,124 confirmed cases outside those categories. Public reports did not define the status of these cases. Potential explanations include pending admission or outcome reconciliation, transfers, and reporting lags, but no category could be assigned without line-list data.

### 3.2 Notification trajectory

The cumulative confirmed count increased approximately seventeen-fold between 27 May and 15 July.

Arithmetic cumulative doubling times lengthened as the denominator accumulated, but this mechanical property of a cumulative curve was not interpreted as evidence of epidemic deceleration.

Notification rates declined gradually across the four June intervals, from 39.7 to 34.1 cases per day, and then increased in July. Excluding the 29 June–1 July interval, the aggregate rate was 35.9 per day during 2–29 June (970 notifications over 27 days; exact counting interval 33.7–38.3) and 47.4 per day during 1–15 July (664 over 14 days; 43.9–51.2). The July-to-June notification-rate ratio was 1.32 (counting-model 95% interval 1.20–1.46).

This increase cannot be interpreted as a direct measure of biological acceleration. WHO noted that expanded surveillance, testing, and diagnostic capacity contributed to rising reported counts, and newly reported cases could include backlogs.[3] However, the reporting-date series showed no sustained decline. WHO also reported that more than 80% of new cases were being detected outside known contact lists, indicating that major transmission chains were still being missed. On 16 July, WHO described the outbreak as having expanded faster during the preceding month than any previous Ebola outbreak.[40] Together, these indicators support continuing epidemiological pressure, while the relative contributions of transmission and ascertainment remain unquantified.

### 3.3 Outcome, contact-tracing, and healthcare-worker indicators

The crude confirmed-case CFR increased from 13.6% on 27 May to 39.0% on 15 July. This rise is compatible with maturation of outcomes among previously active cases, retrospective confirmation of deaths, delayed presentation, preferential detection of severe disease, delayed recovery reporting, and changes in testing or death investigation. It should not be interpreted as a time-resolved measure of clinical virulence.

As of 15 July, 12,693 identified contacts were reported across Ituri, North Kivu, and Tshopo, of whom 10,195 were followed, yielding an overall listed-contact follow-up proportion of 80.3%. Provincial proportions differed substantially: 78.1% in Ituri, 91.7% in North Kivu, and 50.0% in Tshopo.[3] This denominator contains only identified contacts. A high follow-up percentage among listed contacts can coexist with poor epidemic visibility when most incident cases were not listed before detection.

Healthcare-worker infections continued to increase, reaching 119 confirmed infections, 61 recoveries, and 36 deaths by 17 July. This signal indicates repeated exposure in healthcare facilities or the community and is consistent with delayed recognition, inadequate IPC implementation, unsafe specimen or patient handling, and pressure on routine services.[3] WHO had also reported payment difficulties and healthcare-worker strikes during the response, adding operational fragility to already strained treatment and surveillance systems.[41]

### 3.4 Uganda and international events

As of 17 July, Uganda’s official dashboard reported 20 confirmed cases, two confirmed deaths, 18 recoveries, and no current admissions. Fifteen infections were imported and five were locally acquired among contacts and healthcare workers linked to imported cases. No confirmed case had been reported since 21 June, and Uganda began the 42-day countdown toward declaration of the end of the outbreak after discharge of the final patient on 16 July. One probable fatal case remained outside the confirmed denominator.[3,4,42]

Uganda’s confirmed-case CFR was 10% (2/20), compared with 39.0% in the DRC confirmed series. The difference is operationally important but cannot establish a causal treatment effect because Uganda’s denominator was small, cases may have differed in timing and severity, and the probable death was excluded from the confirmed denominator. Uganda’s Ministry of Health reported comprehensive supportive care and medicines administered under compassionate-use protocols, but the individual contribution of experimental products cannot be separated from early detection, specialised isolation, multidisciplinary care, and selection effects.[42]

WHO’s jurisdictional accounting on 17 July comprised 2,145 confirmed cases globally: 2,124 in the DRC, 20 in Uganda, and one in France, with 830 confirmed-case deaths. The two United States workers treated in Germany were diagnosed in the DRC and were already included in the DRC total. The international pattern remained dominated by direct occupational exposure and controlled medical evacuation.[3]

### 3.5 Risk assessment for North Africa

The most plausible introduction pathways for North Africa were commercial aviation, returning humanitarian or occupational personnel, multi-leg travel through African, European, Gulf, or Turkish hubs, and controlled medical evacuation. Published route availability established plausible pathways but did not quantify their magnitude. Morocco had advertised direct Kinshasa–Casablanca service, while Egypt had substantial hub connectivity with East and Central Africa.[13–16] Sudan had a distinct multi-stage overland vulnerability through South Sudan because of conflict-driven mobility and disrupted surveillance, although Ebola’s clinical severity limits prolonged travel after symptom onset.

For Algeria, the principal vulnerability was failure to recognise the first patient in an ordinary public or private healthcare setting. A final departure airport may conceal earlier travel to eastern DRC; triage therefore requires a complete 21-day itinerary and exposure history. Introduction was assessed as very low to low, limited secondary transmission as low following delayed recognition, sustained community transmission as very low under functioning surveillance and IPC, and impact as high because a single missed case could require extensive contact tracing, decontamination, occupational monitoring, and service reorganisation.

### 3.6 Risk assessment for Europe

ECDC estimated approximately one importation per 23,000–24,000 travellers from the principal outbreak region, with a 90% uncertainty interval of one per 13,000–54,000. Under a hypothetical scenario of 100 travellers during 11–25 June, ECDC estimated a 0.45% probability of at least one importation (90% uncertainty interval 0.20%–0.85%). These were separately generated Monte Carlo summaries and should not be forced into exact reciprocal equivalence.[7]

ECDC assessed infection risk for people living in the EU/EEA as very low. France, Belgium, Germany, and the United Kingdom had identifiable route, humanitarian, institutional, or specialised-treatment links that could increase their relative likelihood of encountering an imported or evacuated case, but route availability was not a substitute for origin–destination passenger volumes. Sustained transmission remained very unlikely because infectiousness begins after symptom onset and specialist isolation, diagnostics, and contact tracing are available. The principal failure scenario was an unrecognised symptomatic patient attending multiple facilities or receiving invasive care without appropriate PPE.[1,7–10,17,18]

## 4. Discussion

### 4.1 Principal findings

This analysis found that the publicly available indicators were inconsistent with effective control in eastern DRC at the mid-July cut-off. The outbreak had reached 2,124 confirmed cases across 46 health zones, with a high confirmed-case mortality burden, extensive healthcare-worker infection, and continued geographical spread. Notifications were approximately 32% higher during the first half of July than during the June comparison period. Although reporting-date data cannot separate true incidence from changes in testing and backlog release, the series did not show a sustained decline. Most importantly, WHO reported that more than 80% of new cases were detected outside known contact lists, demonstrating major failure of prospective transmission-chain identification.[3,40]

Uganda provided a contrasting operational signal. After 15 imported and five linked secondary cases, no new confirmed case had been reported since 21 June, the final patient was discharged, and the country entered the 42-day observation period. This pattern shows that cross-border importation and healthcare-associated secondary transmission can occur without progression to sustained community transmission when surveillance, isolation, contact management, and specialised care function effectively.[3,4,42]

For North Africa and Europe, the probability of introduction was low but non-zero, whereas the conditional probability of sustained community transmission was substantially lower. Returning responders, healthcare workers, and household caregivers of an unrecognised case were the principal risk groups. The most important preparedness failure would be delayed recognition in routine clinical care, not failure of thermal or border screening to detect an asymptomatic incubating traveller.

### 4.2 Interpreting notification growth and fatality indicators

The notification-rate analysis improves on simple cumulative doubling-time comparisons because it examines changes over defined intervals. Nevertheless, notification rates remain composite measures of infection, care seeking, alert investigation, specimen transport, laboratory capacity, retrospective death investigation, and reporting. The exact Poisson intervals are therefore deliberately described as counting-model intervals rather than complete epidemiological uncertainty intervals.

The rise in crude confirmed-case CFR should also be interpreted cautiously. Early in an outbreak, many recent cases have unresolved outcomes, producing right truncation. Subsequent confirmation of deaths and delayed recording of recoveries can increase the crude CFR even if the underlying clinical risk is unchanged. Differences between Uganda and the DRC may reflect care pathways and health-system performance, but the available aggregate data do not support causal attribution to individual therapies.

### 4.3 Contact-tracing denominators and hidden transmission

The distinction between follow-up of listed contacts and identification of the contact network is central. An 80.3% follow-up rate among identified contacts may appear reassuring in isolation, yet more than 80% of new cases were reportedly outside existing lists. This combination means that teams were following many known contacts but prospectively identifying only a minority of the chains generating incident cases. Retrospective linkage may later explain some cases, but it does not replace timely listing before symptoms or diagnosis.

Community deaths, delayed presentation, insecurity, population movement, and healthcare-worker infections create mutually reinforcing blind spots. Patients who remain in the community can expose caregivers, transporters, traditional healers, healthcare staff, and burial participants. Insecurity and mistrust delay investigation, while overwhelmed or unpaid staff reduce the capacity to identify and monitor contacts. Consequently, control should be judged using multiple indicators: decline by symptom-onset date, falling community mortality, near-complete alert and death investigation, increasing proportions of cases already known as contacts, fewer healthcare-worker infections, no new affected health zones, shorter onset-to-isolation delays, and absence of unexplained clusters for at least two incubation periods.

### 4.4 International risk and the limits of border measures

An incubating traveller can cross a border while asymptomatic because people are not considered infectious before symptom onset and the incubation period can extend to 21 days. Temperature screening may detect some symptomatic travellers but cannot exclude infection during incubation. Conversely, a recognised symptomatic case can usually be contained because transmission requires direct contact with blood or other body fluids, contaminated materials, or the body of a deceased case rather than ordinary casual proximity.[3,11]

WHO’s temporary recommendations did not support suspension of flights or denial of entry from affected countries. Additional measures should be evidence-based, proportionate, respectful of rights, and notified under the IHR. Indiscriminate restrictions may encourage route concealment, impede responder movement, and intensify stigma. The higher-yield strategy is exposure management before travel, complete itinerary and occupational histories, rapid self-reporting, accessible clinical assessment, protected care, and immediate public-health notification.[20]

### 4.5 Countermeasures and survivor-associated considerations

No vaccine or therapeutic was licensed specifically for BDBV at the cut-off. WHO-sponsored PARTNERS trial enrolment began on 2 July to evaluate MBP134, remdesivir, and their combination. WHO experts also prioritised oral obeldesivir for research as post-exposure prophylaxis and advanced candidate BDBV vaccine platforms.[21–23,34] Ervebo is licensed for disease caused by Ebola virus, not BDBV. Limited nonhuman-primate evidence suggests possible heterologous protection, but WHO considered the evidence insufficient for programmatic BDBV use outside controlled research.[25,37]

With hundreds of survivors, follow-up programmes were epidemiologically relevant. Ebolaviruses can persist in immune-privileged sites and selected body fluids after recovery. Evidence is strongest for Ebola virus rather than BDBV; durations and frequencies should not be assumed equivalent. Survivor care should include post-Ebola clinical support, sexual-health advice and semen testing where recommended, investigation of possible relapse, and anti-stigma communication.[20,24,33]

### 4.6 Strengths and limitations

The principal strength of this study is denominator discipline. Confirmed, probable, and suspected series were kept separate; medical evacuations were not double counted; the unresolved DRC status total was retained as a data gap; and the ECDC Monte Carlo summaries were not treated as exact reciprocals. The analysis also separated introduction, onward transmission, impact, and confidence rather than collapsing them into a single opaque label.

The main limitation is dependence on public aggregate data. Reporting dates do not represent symptom onset, and the analysis could not estimate time-varying reproduction numbers, serial intervals, onset-to-isolation delays, household secondary-attack rates, health-zone-specific growth, or intervention effects. Passenger volumes, traveller occupations, deployment rosters, medevac activity, and country-specific laboratory and referral readiness were incomplete. Country ratings were therefore comparative judgements rather than calibrated probabilities. The analysis was time-stamped and may be superseded by subsequent official reports.

## 5. Operational preparedness implications

### 5.1 Priorities for North Africa and Europe

Preparedness should concentrate on the first clinical encounter. Acute-care systems should obtain a complete 21-day residence, travel, transit, occupational, funeral, healthcare, and body-fluid exposure history from patients with compatible febrile or gastrointestinal illness. The final airport of departure is insufficient because it may conceal earlier presence in an affected health zone.

For a suspected case, systems should immediately place the patient in a designated room, restrict access, use appropriate PPE with observed donning and doffing, avoid unnecessary invasive procedures and movement, notify infection-control and public-health authorities, consult a designated reference laboratory before specimen collection, use validated packaging and transport, and start an exposure log without waiting for laboratory confirmation.

Countries should verify that molecular assays detect BDBV, designate primary and backup laboratories, test after-hours specimen transport, protect routine laboratories from unplanned exposure, and maintain safe waste and decontamination procedures. Simulation exercises should include emergency departments, maternity care, ambulances, laboratories, private facilities, and points of entry. Returning responders should receive exposure-based assessment and 21-day monitoring rather than uniform restrictions.

### 5.2 Algeria-specific priority plan

### 5.3 Indicators requiring immediate reassessment

- Sustained transmission in Kinshasa, Goma, Kisangani, Kampala, or another major international transport hub.
- An imported case in an ordinary commercial traveller without recognised high-risk exposure.
- Multiple international cases without known epidemiological links.
- Sustained transmission in additional neighbouring countries.
- Further increase in the proportion of cases outside known contact lists, deterioration in contact follow-up, or longer laboratory turnaround.
- Continued increase in healthcare-worker infections, major security deterioration, healthcare-worker strikes, or withdrawal of response teams.
- Persistent or increasing community mortality, simultaneous outbreaks that divert capacity, or breakdown of specialist isolation and medical-evacuation arrangements.
- Validated evidence of altered transmission characteristics.

## 6. Conclusion

The 2026 BDBV outbreak was exceptionally large and remained active across eastern DRC at the mid-July data cut-off. Reporting-date notifications were materially higher during the first half of July than during the June comparison period, but public aggregate data could not separate biological transmission from changes in ascertainment. The strongest evidence of inadequate control was not the cumulative count itself, but the combination of more than 80% of new cases being detected outside known contact lists, persistent community mortality, continued healthcare-worker infection, and operational disruption.

Introduction into North Africa or Europe was possible but uncommon. Sustained community transmission remained very unlikely in settings that rapidly recognised and isolated the first case. The decisive preparedness capabilities were complete exposure histories, early clinical suspicion, protected care, validated diagnostics, immediate notification, and effective contact management. Border screening alone could not exclude infection in incubating travellers.

## Data Availability

All data used in this study are publicly available from official sources. The datasets analyzed were obtained from publicly accessible reports and dashboards published by the World Health Organization (WHO), the World Health Organization Regional Office for Africa (WHO AFRO), the World Health Organization Regional Office for the Eastern Mediterranean (WHO EMRO), the European Centre for Disease Prevention and Control (ECDC), Africa CDC, the Uganda Ministry of Health, the UK Health Security Agency (UKHSA), and the U.S. Centers for Disease Control and Prevention (CDC). All data sources are cited in the manuscript and reference list. No new datasets were generated during this study.

## Declarations

### Ethics approval and consent to participate

This study analysed only publicly available aggregate surveillance data and did not involve identifiable human participants, recruitment, intervention, human tissue, or access to confidential records. Formal research-ethics review and consent to participate were therefore not applicable.

### Consent for publication

Not applicable.

### Funding

No external, institutional, or commercial funding was received for this work.

### Competing interests

The authors declare no competing interests.

### Data and code availability

All source data are publicly available through the cited reports. The cumulative counts, intervals, and formulas required to reproduce the derived notification rates and rate ratio are provided in the main text and Appendix A. No individual-level or confidential data were accessed.

### Author contributions

O.W.B. conceived and designed the study; identified and reviewed sources; extracted and reconciled aggregate data; performed the quantitative analysis; developed the risk-assessment framework; interpreted the findings; and drafted the manuscript. M.Y.K. provided scientific supervision, reviewed the epidemiological methods and interpretation, and critically revised the manuscript. Both authors approved the final manuscript and accept responsibility for its content.

### Use of generative artificial intelligence

During drafting and revision, generative artificial-intelligence tools (Anthropic Claude and OpenAI ChatGPT) were used under the authors’ supervision for language editing, structural refinement, and assistance in checking arithmetic, statistical presentation, and references. The authors independently reviewed the full manuscript, verified substantive claims and calculations against the cited sources, and accept full responsibility for the final content. No artificial-intelligence system is listed as an author.

## Acknowledgements

None.

## Appendix A. Reproducibility of the notification-rate analysis

The interval analysis used the cumulative confirmed DRC counts shown in Table 3. For an interval bounded by reports at dates d0 and d1, the number of newly notified cases was C1 ™ C0 and the average daily rate was (C1 ™ C0)/(d1 ™ d0). Exact Garwood limits for the Poisson count were divided by the interval duration. The aggregate June comparison used 970 notifications during 27 days from 2–29 June; the July comparison used 664 notifications during 14 days from 1–15 July. The resulting rate ratio was (664/14)/(970/27) = 1.3202. The log-scale standard error was √(1/664 + 1/970), yielding a 95% counting-model interval of 1.1961–1.4572, reported as 1.20–1.46.

**Table 1.** Operational definitions for introduction likelihood.

| Category | Operational reference class over 4–8 weeks |
| --- | --- |
| Very low | Biologically possible but requiring an uncommon convergence of exposure, travel, and detection failure; no case expected under ordinary conditions. |
| Low | A documented and plausible pathway exists; sporadic introduction would not be surprising but is not expected as a routine event. |
| Moderate | Recurrent pathways, active regional transmission, or repeated exportations make introduction a credible event that preparedness systems should expect to encounter. |
| High / very high | Active local or immediately adjacent transmission makes further cases expected without substantial control. |

**Note:** Impact and confidence were graded separately. Adjacent-category ranges indicate genuine uncertainty near a boundary, not a distinct category.

**Table 2.** Confirmed DRC cases and deaths by province as of 15 July 2026.

| Province | Confirmed cases | Deaths | Affected health zones |
| --- | --- | --- | --- |
| Ituri | 1,904 | 692 | 27 of 36 |
| North Kivu | 199 | 119 | 11 of 34 |
| South Kivu | 3 | 1 | 1 of 34 |
| Haut-Uele | 14 | 13 | 4 of 13 |
| Tshopo | 4 | 3 | 3 of 23 |
| Total | 2,124 | 828 | 46 of 140 |

**Note:** Source: WHO and ECDC.[1,3]

**Table 3.**
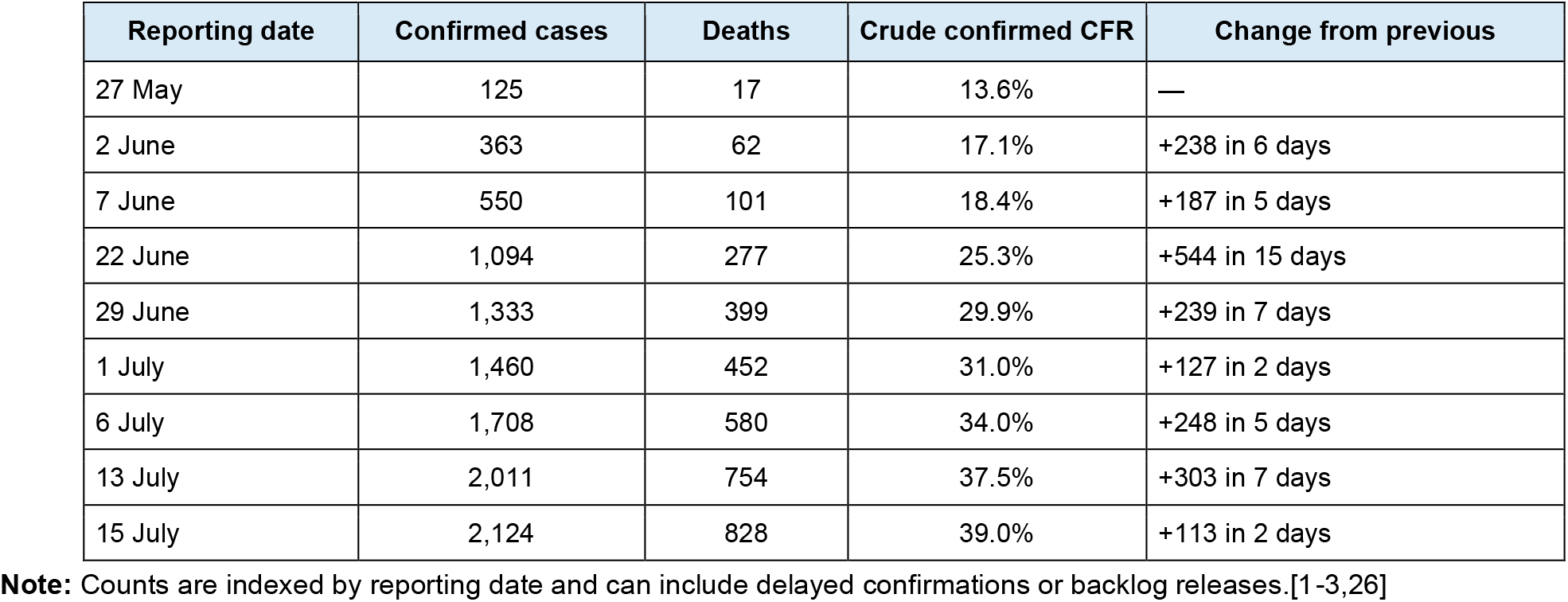
Selected cumulative confirmed-case milestones in the DRC.

**Table 4.** Interval-specific DRC confirmed-case notification rates.

| Interval | New notifications | Days | Mean/day | Exact Poisson 95% counting interval |
| --- | --- | --- | --- | --- |
| 27 May–2 June | 238 | 6 | 39.7 | 34.8–45.0 |
| 2–7 June | 187 | 5 | 37.4 | 32.2–43.2 |
| 7–22 June | 544 | 15 | 36.3 | 33.3–39.4 |
| 22–29 June | 239 | 7 | 34.1 | 30.0–38.8 |
| 29 June–1 July | 127 | 2 | 63.5 | 52.9–75.6 |
| 1–6 July | 248 | 5 | 49.6 | 43.6–56.2 |
| 6–13 July | 303 | 7 | 43.3 | 38.5–48.4 |
| 13–15 July | 113 | 2 | 56.5 | 46.6–67.9 |

**Note:** Exact Garwood intervals describe counting variation only and exclude surveillance-system uncertainty.

**Table 5.** Comparative rapid risk assessment for North Africa.

| Country | Introduction | Onward transmission after one importation | Impact if recognition delayed | Confidence | Rationale |
| --- | --- | --- | --- | --- | --- |
| Morocco | Low | Very low to low | High | Moderate | Direct/hub connectivity increases relative exposure; prompt recognition should keep onward spread limited. |
| Egypt | Low | Very low to low | High | Moderate | Large aviation hub and East/Central African links; delayed recognition in dense healthcare settings would have substantial consequences. |
| Algeria | Very low to low | Very low to low | High | Moderate | Introduction is more plausible through indirect travel or returning personnel than direct outbreak-area traffic. |
| Tunisia | Very low to low | Very low | Moderate to high | Moderate | Mainly connecting-flight and occupational pathways; impact depends on referral speed. |
| Libya | Very low | Low, conditional on a missed case | Very high | Low | Arrival is uncommon, but fragmented services and insecurity could amplify a missed case. |
| Sudan | Low | Low to moderate, conditional | Very high | Low | Regional mobility through South Sudan, conflict, displacement, and surveillance disruption increase uncertainty. |
| Mauritania | Very low | Very low to low | Moderate to high | Low to moderate | Limited direct exposure; impact is governed mainly by laboratory access and referral speed. |

**Note:** Ratings are comparative judgements, not calibrated probabilities. Passenger-volume and confidential readiness data were unavailable.

**Table 6.** Europe-specific rapid risk assessment.

| Population or setting | Exposure / importation | Onward transmission | Impact | Confidence | Assessment |
| --- | --- | --- | --- | --- | --- |
| General EU/EEA public | Very low | Very low | Moderate | High | Population risk remains very low; impact rises if recognition is substantially delayed. |
| Travellers without direct exposure | Very low | Very low | Moderate | High | Routine advice and symptom awareness are appropriate. |
| Returning responders | Low, but above ordinary travellers | Low if exposure-based monitoring functions | High | Moderate | Priority group for structured exposure assessment and monitoring. |
| Household contacts of an unrecognised case | Conditional on importation | Low to moderate without precautions | High | Moderate | Rapid identification and protection from body-fluid exposure are essential. |
| First-contact healthcare workers | Rare exposure, high consequence | Moderate if invasive care precedes recognition | Very high | High | Highest preparedness priority in routine care. |
| Specialist isolation / medevac teams | Repeated but controlled exposure | Very low with protocols | High | High | Maintain drills, PPE competence, and staffing resilience. |

**Note:** The table separates population likelihood from consequences for specific occupational or household groups.

**Table 7.**
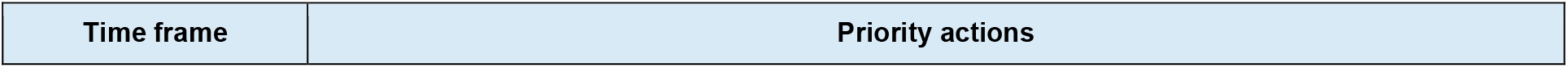
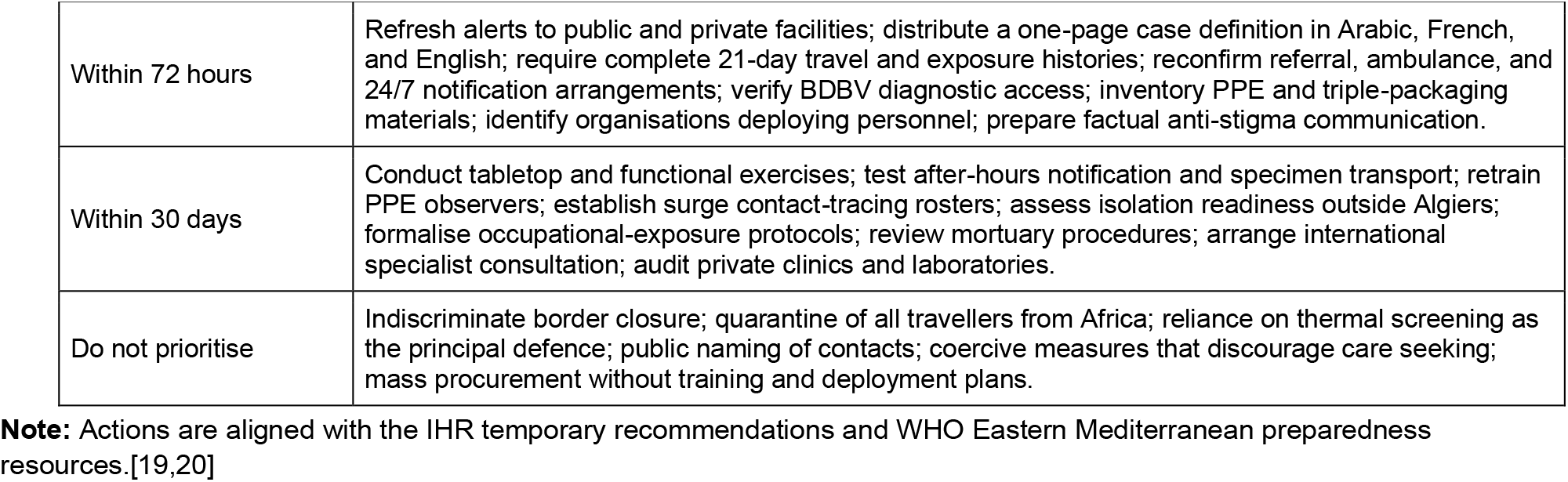
Priority preparedness actions for Algeria.

| Time frame | Priority actions |
| --- | --- |
| Within 72 hours | Refresh alerts to public and private facilities; distribute a one-page case definition in Arabic, French, and English; require complete 21-day travel and exposure histories; reconfirm referral, ambulance, and 24/7 notification arrangements; verify BDBV diagnostic access; inventory PPE and triple-packaging materials; identify organisations deploying personnel; prepare factual anti-stigma communication. |
| Within 30 days | Conduct tabletop and functional exercises; test after-hours notification and specimen transport; retrain PPE observers; establish surge contact-tracing rosters; assess isolation readiness outside Algiers; formalise occupational-exposure protocols; review mortuary procedures; arrange international specialist consultation; audit private clinics and laboratories. |
| Do not prioritise | Indiscriminate border closure; quarantine of all travellers from Africa; reliance on thermal screening as the principal defence; public naming of contacts; coercive measures that discourage care seeking; mass procurement without training and deployment plans. |

**Note:** Actions are aligned with the IHR temporary recommendations and WHO Eastern Mediterranean preparedness resources.[19,20]

These calculations quantify variation under a Poisson counting model only. They do not adjust for delayed confirmations, backlogs, changing diagnostic capacity, under-ascertainment, serial correlation, or other surveillance-process effects. Therefore, they should not be interpreted as confidence limits for the underlying infection incidence.

## Notes

### Competing Interest Statement

The authors have declared no competing interest.

### Author Declarations

The study used only openly available, aggregated human surveillance data obtained from official public sources, including the World Health Organization (WHO) Disease Outbreak News and Rapid Risk Assessments, the European Centre for Disease Prevention and Control (ECDC), Africa CDC, the Uganda Ministry of Health Ebola Dashboard, the WHO Regional Office for Africa (AFRO), the WHO Regional Office for the Eastern Mediterranean (EMRO), the UK Health Security Agency (UKHSA), and the U.S. Centers for Disease Control and Prevention (CDC). All data were publicly accessible before the study was initiated. No individual-level patient data, restricted-access datasets, or confidential information were used.

## References

1. European Centre for Disease Prevention and Control. Ebola disease outbreak in the Democratic Republic of the Congo and Uganda. Updated 17 July 2026. https://www.ecdc.europa.eu/en/ebola-outbreak-democratic-republic-congo-and-uganda

2. European Centre for Disease Prevention and Control. Communicable Disease Threats Report, week 29, 12–18 July 2026. https://www.ecdc.europa.eu/sites/default/files/documents/communicable-disease-threats-report-week-29-2026.pdf

3. World Health Organization. Disease Outbreak News: Ebola disease caused by Bundibugyo virus—Democratic Republic of the Congo and Uganda (DON613). 17 July 2026. https://www.who.int/emergencies/disease-outbreak-news/item/2026-DON613

4. Uganda Ministry of Health. Ebola outbreak dashboard and updates. https://evd-daily.health.go.ug/

5. World Health Organization. Rapid risk assessment: Bundibugyo virus disease outbreak in the Democratic Republic of the Congo and Uganda, version 3. 6 June 2026. https://cdn.who.int/media/docs/default-source/_sage-2026/who-rapid-risk-assessment-ebola-bundibugyo-virus-disease--democratic-republic-of-the-congo--uganda--and-countries-with-land-borders-adjoining-countries-with-documented-bdbv-detection_v3.pdf

6. WHO Regional Office for Africa. Outbreak information and technical guidance: Ebola disease, DRC 2026.https://www.afro.who.int/health-topics/ebola-disease/outbreak-drc-26

7. European Centre for Disease Prevention and Control. Estimation of the importation risk of Bundibugyo virus into the EU/EEA in June 2026. 15 June 2026. https://www.ecdc.europa.eu/en/publications-data/estimation-importation-risk-bundibugyo-virus-eueea-june-2026

8. European Centre for Disease Prevention and Control. Preparedness and response for imported cases of Ebola disease into an EU/EEA country. Updated 10 July 2026. https://www.ecdc.europa.eu/en/publications-data/preparedness-and-response-imported-cases-ebola-disease-eueea-country-updated-10

9. WHO Regional Office for Europe. Ebola outbreak in Africa: preparedness in the WHO European Region. https://www.who.int/europe/emergencies/situations/ebola-outbreak-in-africa-preparedness-in-the-who-european-region

10. UK Health Security Agency. Ebola disease outbreak in the Democratic Republic of the Congo and Uganda: Bundibugyo virus. https://www.gov.uk/guidance/ebola-disease-outbreak-in-the-democratic-republic-of-the-congo-and-uganda-bundibugyo-virus

11. World Health Organization. Ebola disease fact sheet. Updated 24 April 2025. https://www.who.int/news-room/fact-sheets/detail/ebola-disease

12. WHO Regional Office for Africa. Bundibugyo Ebola Virus Continental Preparedness and Response Plan, June–November 2026. https://www.afro.who.int/publications/bundibugyo-ebola-virus-continental-preparedness-and-response-plan-june-november-2026

13. Royal Air Maroc. Flights from Kinshasa to Casablanca. https://www.royalairmaroc.com/en/flights-from-kinshasa-to-casablanca

14. Royal Air Maroc. Flights from Casablanca to Kinshasa. https://www.royalairmaroc.com/en/flights-from-casablanca-to-kinshasa

15. EgyptAir. Kinshasa–Cairo route information. https://www.egyptair.com/en/Plan/special-offers/Pages/Kinshasa-Cairo.aspx

16. EgyptAir. Network and timetable information. https://www.egyptair.com/en

17. UK Health Security Agency. Outbreaks under monitoring: week 28, 2026. https://www.gov.uk/government/publications/outbreaks-under-monitoring-in-2026/outbreaks-under-monitoring-week-28-week-ending-12-july-2026

18. US Centers for Disease Control and Prevention. Ebola outbreak: current situation. https://www.cdc.gov/ebola/situation-summary/index.html

19. WHO Regional Office for the Eastern Mediterranean. Ebola preparedness resources. https://www.emro.who.int/health-topics/ebola/ebola.html

20. World Health Organization. First meeting of the IHR Emergency Committee regarding the epidemic of Ebola Bundibugyo virus disease in the Democratic Republic of the Congo and Uganda—temporary recommendations. 22 May 2026. https://www.who.int/news/item/22-05-2026-first-meeting-of-the-ihr-emergency-committee-regarding-the-epidemic-of-ebola-bundibugyo-virus-disease-in-the-democratic-republic-of-the-congo-and-uganda-2026-temporary-recommendations

21. World Health Organization. Patient enrolment begins in a scientific trial to identify the first effective treatments for Bundibugyo virus disease. 2 July 2026. https://www.who.int/news/item/02-07-2026-patient-enrolment-begins-in-a-scientific-trial-to-identify-the-first-effective-treatments-for-bundibugyo-virus-disease

22. World Health Organization. WHO adds first diagnostic test for Ebola Bundibugyo virus to its Emergency Use Listing. 2 July 2026. https://www.who.int/news/item/02-07-2026-who-adds-first-diagnostic-test-for-ebola-bundibugyo-virus-to-its-emergency-use-listing

23. World Health Organization. WHO target product profile for Bundibugyo virus disease vaccines. 14 July 2026.https://www.who.int/publications/i/item/B09835

24. World Health Organization. Clinical care for survivors of Ebola virus disease: interim guidance. 2016.https://iris.who.int/bitstreams/e237ca3b-124c-444e-8f1e-a0d95480ab54/download

25. World Health Organization. Emergency guidance on the use of licensed Ebola virus vaccine during Bundibugyo virus disease outbreaks. 28 May 2026. https://www.who.int/publications/i/item/B09772

26. World Health Organization. Disease Outbreak News: Ebola disease caused by Bundibugyo virus—Democratic Republic of the Congo (DON603). 21 May 2026. https://www.who.int/emergencies/disease-outbreak-news/item/2026-DON603

27. US Centers for Disease Control and Prevention. CDC statement on Ebola outbreak in the Democratic Republic of the Congo and Uganda. 10 July 2026. https://www.cdc.gov/media/releases/2026/cdc-ebola-outbreak-in-democratic-republic-of-congo-and-uganda.html

28. Zomahoun DL, Boyd MA, Honein MA, et al. Outbreak of Ebola disease caused by Bundibugyo virus—Democratic Republic of the Congo and Uganda, May 2026. MMWR Morb Mortal Wkly Rep. 2026;75:293–294. https://www.cdc.gov/mmwr/volumes/75/wr/mm7522e3.htm

29. Towner JS, Sealy TK, Khristova ML, et al. Newly discovered Ebola virus associated with hemorrhagic fever outbreak in Uganda. PLoS Pathog. 2008;4:e1000212. https://pmc.ncbi.nlm.nih.gov/articles/PMC2581435/

30. MacNeil A, Farnon EC, Morgan OW, et al. Proportion of deaths and clinical features in Bundibugyo Ebola virus infection, Uganda. Emerg Infect Dis. 2010;16:1969–1972. https://pmc.ncbi.nlm.nih.gov/articles/PMC3294552/

31. Kratz T, Roddy P, Tshomba Oloma A, et al. Ebola virus disease outbreak in Isiro, Democratic Republic of the Congo, 2012: signs and symptoms, management and outcomes. PLoS One. 2015;10:e0129333. https://pmc.ncbi.nlm.nih.gov/articles/PMC4479598/

32. US Centers for Disease Control and Prevention. History of Ebola outbreaks. Updated 29 May 2026. https://www.cdc.gov/ebola/outbreaks/index.html

33. Mukadi-Bamuleka D, et al. Fatal meningoencephalitis associated with Ebola virus persistence. Lancet Microbe. 2024. https://www.thelancet.com/journals/lanmic/article/PIIS2666-5247(24)00137-X/fulltext

34. World Health Organization. Experts convened by WHO advise on candidate treatments and vaccines for Ebola disease caused by Bundibugyo virus. 28 May 2026. https://www.who.int/news/item/28-05-2026-experts-convened-by-who-advise-on-candidate-treatments-and-vaccines-for-ebola-disease-caused-by-bundibugyo-virus

35. MacNeil A, Farnon EC, Morgan OW, et al. Filovirus outbreak detection and surveillance: lessons from Bundibugyo. J Infect Dis. 2011;204(Suppl 3):S761–S767. doi:10.1093/infdis/jir294. https://pubmed.ncbi.nlm.nih.gov/21987748/

36. Hulseberg CE, Kumar R, Di Paola N, et al. Molecular analysis of the 2012 Bundibugyo virus disease outbreak. Cell Rep Med. 2021;2:100351. doi:10.1016/j.xcrm.2021.100351. https://pmc.ncbi.nlm.nih.gov/articles/PMC8385243/

37. Falzarano D, Feldmann F, Grolla A, et al. Single immunization with a monovalent vesicular stomatitis virus-based vaccine protects nonhuman primates against heterologous challenge with Bundibugyo ebolavirus. J Infect Dis. 2011;204(Suppl 3):S1082–S1089. doi:10.1093/infdis/jir350. https://pmc.ncbi.nlm.nih.gov/articles/PMC3189995/

38. WHO Regional Office for Africa. Ebola virus disease—Democratic Republic of the Congo: historical outbreak summary. https://www.afro.who.int/news/ebola-virus-disease-democratic-republic-congo

39. Africa CDC Knowledge Hub. Key facts about the Bundibugyo virus outbreak. https://khub.africacdc.org/records/resource/key-facts-about-the-bundibugyo-virus-outbreak

40. World Health Organization. WHO Director-General’s opening remarks at the media briefing. 16 July 2026. https://www.who.int/news-room/speeches/item/who-director-general-s-opening-remarks-at-the-media-briefing---16-july-2026

41. United Nations Office at Geneva. UN Geneva Press Briefing. 7 July 2026. https://www.ungeneva.org/en/news-media/press-briefing/2026/07/un-geneva-press-briefing-0

42. Uganda Ministry of Health. Uganda discharges last Ebola patient, begins 42-day countdown towards declaring the outbreak over. 16 July 2026. https://health.go.ug/uganda-discharges-last-ebola-patient-begins-42-day-countdown-towards-declaring-the-outbreak-over/

